# Theta and gamma connectivity is linked with affective and cognitive symptoms in Parkinson’s disease

**DOI:** 10.1101/2020.07.22.20158469

**Authors:** Kartik K. Iyer, Tiffany R. Au, Anthony J. Angwin, David A. Copland, Nadeeka N. Dissanayaka

**Affiliations:** UQ Centre for Clinical Research, Faculty of Medicine, The University of Queensland, Royal Brisbane & Women’s Hospital, Herston QLD 4029, Brisbane, Australia.; QIMR Berghofer Medical Research Institute, Clinical Brain Networks group, Australia.; Department of Neurology, Royal Brisbane & Women’s Hospital, Herston QLD 4029, Brisbane, Australia; School of Psychology, The University of Queensland, St Lucia, QLD 4067, Brisbane, Australia; School of Health & Rehabilitation Sciences, The University of Queensland, St Lucia, QLD 4067, Brisbane, Australia

**Author notes:** **Corresponding Author** Dr Nadeeka Dissanayaka, **Postal address:** University of Queensland, UQ Centre for Clinical Research, Building 71/918 Royal Brisbane & Women’s Hospital, Herston QLD, 4029, Brisbane, Australia.

**Keywords:** brain connectivity, Parkinson’s disease, affective and cognitive symptoms, cortical oscillations

## Abstract

**Background:** The progression of Parkinson’s disease (PD) can often exacerbate symptoms of depression, anxiety, and/or cognitive impairment. In this study, we explore the possibility that multiple brain network responses are associated with symptoms of depression, anxiety and cognitive impairment in PD. This association is likely to provide insights into a single multivariate relationship, where common affective symptoms occurring in PD cohorts are related with alterations to electrophysiological response.

**Methods:** 70 PD patients and 21 healthy age-matched controls (HC) participated in a high-density electroencephalography (EEG) study. Functional connectivity differences between PD and HC groups of oscillatory activity at rest and during completion of an emotion-cognition task were examined to identify key brain oscillatory activities. A canonical correlation analysis (CCA) was applied to identify a putative multivariate relationship between connectivity patterns and affective symptoms in PD groups.

**Results:** A CCA analysis identified a single mode of co-variation linking theta and gamma connectivity with affective symptoms in PD groups. Increases in frontotemporal gamma, frontal and parietal theta connectivity were related with increased anxiety and cognitive impairment. Decreases in temporal region theta and frontoparietal gamma connectivity were associated with higher depression ratings and PD patient age.

**Limitations:** This study only reports on optimal dosage of dopaminergic treatment (‘on’ state) in PD and didn’t investigate at “off” medication.

**Conclusions:** Theta and gamma connectivity during rest and task-states are linked to affective and cognitive symptoms within fronto-temporo-parietal networks, suggesting a potential assessment avenue for understanding brain-behavior associations in PD with electrophysiological task paradigms.

## 1.0 Introduction

Parkinson’s disease (PD) is a well characterised neurodegenerative condition that is predominantly known to influence the motor system (Poewe et al., 2017). PD also results in the heterogeneous manifestation of common non-motor symptoms including anxiety, depression and impaired cognition (Aarsland et al., 2017; Broen et al., 2016; Pellicano et al., 2015; Poletti et al., 2012). Anxiety and depression can also present at the prodromal stage of PD (Postuma and Berg, 2016). Individuals with PD are debilitated by these affective and cognitive symptoms, which interacts with the severity of the disease, impacting their mental health and contributing to a poor quality of life (Ehgoetz Martens and Shine, 2018).

The manifestation of affective and cognitive symptoms in PD have been associated with significant changes to whole-brain functional and structural connectivity (Cerasa et al., 2016; Lanskey et al., 2018; Prell, 2018). Recent neuroimaging work has suggested that such neuropsychiatric manifestations of PD are supported by altered functional connectivity between regions defining default mode frontoparietal, and limbic networks (Amboni et al., 2015; Bell et al., 2015; Hassan et al., 2017; Hu et al., 2015; O’Callaghan et al., 2016; Wee et al., 2016). These results are in line with structural neuroimaging findings which suggest that cognitive impairment in PD is linked to white matter alterations in frontal and parietal regions (Hall et al., 2016). More generally, studies have also put forward the possible use of affective and cognitive symptoms to parse the heterogeneity of PD patients into more homogenous sub-groups (Ehgoetz Martens and Shine, 2018; Sauerbier et al., 2016).

Several studies have also examined how specific cognitive and affective symptoms in PD groups alter spatiotemporal patterns of cortical activity. Previous literature has shown that higher delta/ theta power in EEG studies, and lower beta band power in MEG studies are best predictors of cognitive decline, leading to conversion of PD dementia (Boon et al., 2019; Geraedts et al., 2018). Alpha and theta oscillations have been linked to apathy and reward based movement, and theta oscillations have been linked to impulse control disorders (Bockova and Rektor, 2019; Zhu et al., 2019). Alterations in alpha and theta rhythms have also known to linked with altered emotional capacity in persons with depression (Fernandez-Palleiro et al., 2020). These studies highlight that observable changes to specific oscillatory frequencies that present within cortical activity patterns are useful dynamic representations of how affective and cognitive symptoms alter brain network response in Parkinson’s populations.

Multivariate techniques including canonical correlation analysis (CCA) are emerging as a powerful tool to interrogate links between brain activity and behaviour (Drysdale et al., 2017; Smith et al., 2015; Xia et al., 2018). Specifically, CCA analyses can be used to assess whether a putative link between brain activity and symptoms exists along single or multiple dimensions (Iyer et al., 2019; Lin et al., 2018). This approach may provide key evidence of neural processes underpinning affective and cognitive symptoms in PD. Such evidence could inform current clinical management and response to ongoing interventions in PD patients who have mood and cognitive problems.

In this study, we put forward the hypotheses that brain connectivity, as examined via oscillatory patterns observed during an electrophysiological experiment, will significantly differ in PD groups with affective symptoms in comparison with healthy age-matched controls. Specifically, differences in fronto-temporo-parietal network response of oscillatory patterns will significantly be altered during resting-states and during completion of a cognitive task. To address our study aims, we measured individual changes to oscillatory brain activity in PD patients using high-density electroencephalography (HD-EEG) under two conditions: (i) resting-state; and (ii) during completion of a simple affective priming task (Dissanayaka et al., 2017a; Dissanayaka et al., 2019). Within these settings, our study also explores whether brain network changes found within PD groups at rest and during cognitive states, can be explained by a common multivariate association that ties together electrophysiological response with symptoms of anxiety, depression and cognitive impairment.

## 2.0 Methods

### 2.1 Subjects

The study recruited 102 right handed PD patients diagnosed according to the United Kingdom Brain Bank criteria (Hughes et al., 1992) by registered neurologists. The following exclusion criteria were applied to ascertain our final sample size: (i) withdrawals from the study (*n*=6), (ii) signs of dementia (*n*=10) as identified by treating neurologists based on clinical impression or scoring less than 24 in the brief Standardised Mini-Mental Examination (SMMSE) (Molloy et al., 1991), (iii) severe right hand tremor that interfered with EEG recordings (*n*=1), (iv) poor segment count (*n*=7), and (v) EEG/experimental error (*n*=8). Following this exclusion criteria, our final sample for this study comprised 70 PD patients. We also included 21 healthy age-matched controls. Demographic and clinical characteristics are shown in **Table 1**.

**Table 1:** Demographic and clinical characteristics of Parkinson’s disease (PD) participants compared to healthy older adults including sample size, age and gender distribution, with mean and standard deviation (SD). No significant differences were found in age (t-test) and gender (Chi square test).

| Characteristic | PD | Healthy controls |
| --- | --- | --- |
| *Demographic* |  |  |
| Sample size ($n$) | 70 | 21 |
| Age | 65.60 (8.89) | 69.76 (5.97) |
| Gender (Males/Females) | 42/28 | 12/9 |
| *Symptom measures* |  |  |
| SMMSE | 28.57 (1.66) | 29.24 (0.94) |
| PD-CRS | 97.41 (14.37) | 99.67 (13.37) |
| Mild Cognitive Impairment based on PD-CRS scores (Yes/No) | 11/59 | n/a |
| HAM-A | 7.93 (8.70)** | 0.71 (1.62) |
| HAM-D | 7.69 (7.60)** | 0.81 (1.44) |
| Current Mini-Plus DSM-IV diagnosis of an anxiety disorder (Yes/No) | 40/30 | 0/21 |
| Current Mini-Plus DSM-IV diagnosis of a depressive disorder (Yes/No) | 11/59 | 0/21 |
| UPDRS-III | 23.23 (8.61) | n/a |
| Hoehn & Yahr | 2.22 (0.54) | n/a |
| Levodopa Equivalent Dose | 641.72 (439.79) | n/a |

\*\*Unpaired t-tests for these assessments suggest that PD group is significantly different to healthy controls at $p_{FDR} < 0.001$ . “—” indicates non-significance.
Symptom measures used: SMMSE: Standardised Mini-Mental State Examination – cognitive assessment; PD-CRS: Parkinson’s Disease Cognitive Rating – cognitive assessment for PD patients; HAM-A: Hamilton Anxiety Rating scale – anxiety rating; HAM-D: Hamilton Depression Rating scale – depression rating; UPDRS-III: Unified Parkinson’s Disease Rating Scale – motor examination.

Furthermore, out of 70 PD patients all but 7 PD patients were taking levodopa replacement therapy. Of the 7 PD patients, 3 PD patients reported taking dopamine agonists only, and 4 PD patients reported not taking any PD medication at all. Those participants who were taking PD medication, participated in the study during dopaminergic “on” states (i.e. optimal functioning of levodopa equivalent daily dose (LEDD) in these participants).

Ethical approval was granted by the Royal Brisbane and Women’s Hospital and the University of Queensland human research ethics committees.

### 2.2 Symptom measures

In this study, we used a standardized and validated test battery to evaluate motor and non-motor symptoms in persons with PD. Motor symptom severity of PD was assessed using the Movement Disorders Society Unified Parkinson’s Disease Rating Scale and Hoehn and Yahr staging (see table 1) (Goetz et al., 2007). We used the PD Cognitive Rating Scale (PD-CRS), a clinically recommended measure of cognitive impairment in PD (Pagonabarraga et al., 2008), where we modified this assessment for our analyses so that higher scores represent higher impairment (PD-CRS = maximum score of 134 minus individual PD-CRS score). The PD-CRS measures global cognition in PD with strong construct validity, high test□retest reliability, inter□rater reliability (Cronbach’s α = 0.85). Using a validated cut-off score of 81 in the PD-CRS, the sample was dichotomised into persons with MCI (n=11) and without MCI (n=59) (table 1) (Fernandez de Bobadilla et al., 2013). We also assessed anxiety and depression using the Hamilton Anxiety Rating Scale (HAM-A) (Hamilton, 1959) and Hamilton Depression Rating Scale (HAM-D) (Hamilton, 1960), respectively. For healthy controls, only the HAM-A, HAM-D and PD-CRS rating scales were recorded. Patients with PD had significantly higher HAM-A and HAM-D scores compared to healthy controls. Patients with PD also received a Diagnostic and Statistical Manual Edition IV (DSM-IV) evaluation for anxiety and depressive disorders diagnosis using the Mini International Neuropsychiatric Inventory (Mini-Plus). There were 40 PD patients with anxiety and 11 with depressive disorders. All assessments were conducted within a 2-week period of our EEG experiments (**Table 1)**.

### 2.3 Experimental paradigm

All our participants performed an automatic affective priming task. The word pairs used in this task were constructed from an established affective norms database for English words (ANEW) (Bradley and Lang, 1999). We did not include positive words due to the relatively small number of positively valanced words in the ANEW database, and to minimize the complexity of switching between task conditions for PD patients. The paradigm consisted of a sequence of negatively valanced (rating < 3) and neutral valanced (ratings 4-6) words which were paired to construct a prime word and target word in congruent and incongruent conditions respectively. In the congruent condition, prime-target word pairs consisted of negative-negative or neutral-neutral pairs, whereas our incongruent condition consisted of negative-neutral or neutral-negative pairs, thus totalling to 4 trial conditions. Each condition consisted of 40 trials (e.g. 40 neutral-negative pairs) for a total of 160 trials. All were presented in random order in 4 blocks (40 trials per block) with breaks in between each block to avoid possible fatigue or effects on attentional resources. Using E-PRIME 2.0 software, each experimental trial commenced with presentation of a fixation cross (500ms), followed by a prime-target word pair with a short stimulus onset asynchrony of 250ms to initiate automatic processes (i.e. a jitter). Participants evaluated the target word (i.e., negative or neutral) by pressing the relevant response button. A short practice task was conducted prior to the start of the paradigm. **Figure 1** summarizes the task paradigm employed in the study during acquisition of high-density electroencephalography (HD-EEG).

**Figure 1:**
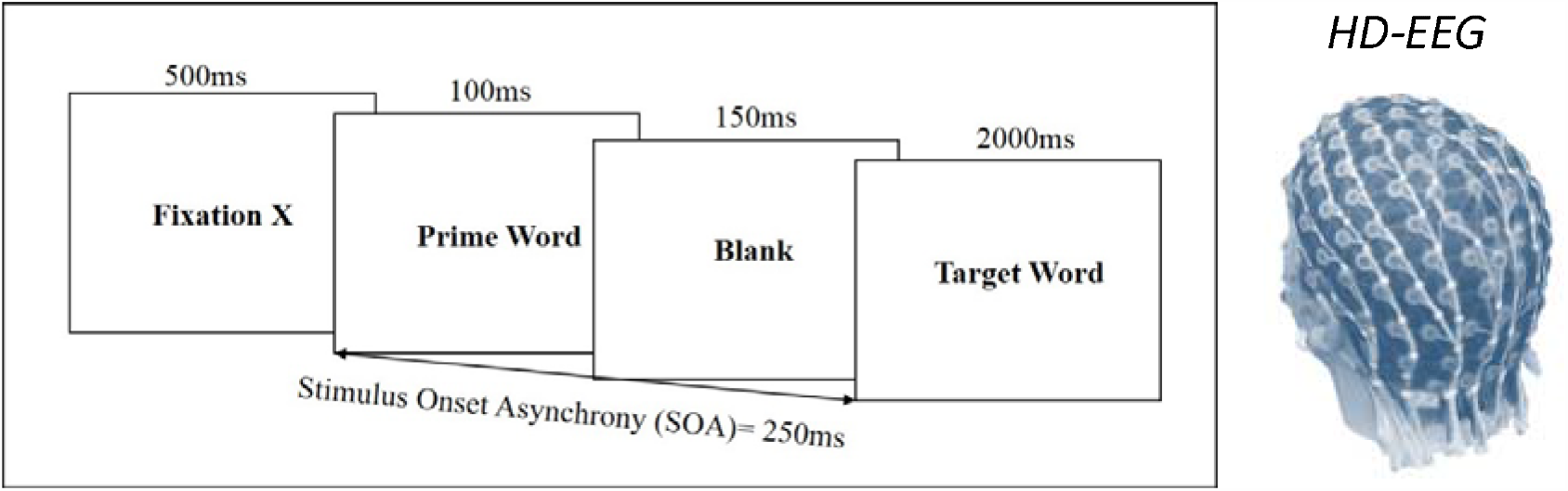
Affective priming task design as measured during HD-EEG recording (image adapted from Dissanayaka et al. (2017a)). All participants were asked to evaluate the valence of the target word with a button press while Event Related Potentials (ERP) via HD-EEG acquisition was recorded simultaneously. (HD-EEG image adapted from Taberna et al. (2019))

### 2.4 Data acquisition and pre-processing

We used an EGI 300 Geodesic EEG system (GES 300, 128-channel system) to acquire HD-EEG. All data was acquired at a sampling rate of 500 Hz. We recorded EEG data under two experimental conditions: (i) resting-state with eyes closed for 3 minutes; and (ii) during performance of an automatic affective priming task. Following data acquisition in both conditions, EEG data was filtered at a lowpass of 30 Hz and a high pass of 0.1 Hz and then downsampled to 250 Hz. Electro-ocular artefacts were monitored by electrodes positioned at the outer canthi of the eye and at infra and supra orbital locations (Hill et al., 2005). Detected electro-ocular artefacts including eye blinks (i.e. defined as >140μV) and eye movements (defined as >55μV), and bad channels (i.e. defined as >200μV for the entire segment) were discarded from the analysis. After artefacts were discarded and bad channels were appropriately replaced, more than 65% of the trials remained for each condition and were included in the analysis. **Table 2** shows the means and standard deviations of the number of trials included in the EEG analysis.

**Table 2:** Mean and standard deviations of the number of trials included in the EEG analysis for Parkinson’s disease (PD) patients and Controls.

| Condition | PD (n=70) | Controls (n=21) |
| --- | --- | --- |
|  | Mean (SD) |  |
| Negative-Negative | 27.86 (7.08) | 29.24 (6.85) |
| Negative-Neutral | 30.70 (5.42) | 30.57 (4.92) |
| Neutral-Negative | 27.09 (7.72) | 28.81 (7.89) |
| Neutral-Neutral | 31.46 (5.92) | 30.81 (5.29) |

Our task-state EEG data considered all prime-target word pairs of Event Related Potentials (ERP) data following baseline correction at −100 ms pre-prime stimulus to 0 ms, relative to the onset of the prime. Baseline corrected ERP windows from 0 to 1000 ms post-target stimulus were then used for synchrony analysis, as this captures the main event-related cortical activity observed with the task (Dissanayaka et al., 2017a; Dissanayaka et al., 2019). Only correct trial responses with reaction times within 2000 ms were included for final analysis. The number of correct trials did not significantly differ across PD and HC groups across congruous and incongruous stimuli.

Resting-state EEG was epoched into 2-second non-overlapping windows to provide reliable estimates for our synchrony measures, described below. An epoch length of 2-seconds was chosen on basis of reliability in deriving phase lag correlations from source-level data (source reconstruction, PLI, described further below) (Fraschini et al., 2016) and availability of epochs (i.e. 3 minutes of data = 90 epochs).

For EEG data in both experimental conditions, source reconstruction was performed using SPM12. Cortical source densities were obtained for all subjects across all experimental conditions and anatomically mapped to a canonical head model comprising 8196 vertices based on the MNI brain as provided by SPM (Litvak et al., 2011). Forward model computation was then performed using a boundary element model (BEM) composed on three layers (scalp, outer skull, inner skull) using a Bayesian localization approach involving multiple sparse priors (MSP). Following reconstruction, we next extracted 68 regional cortical time-series, in resting and task state EEG epochs, via source inversion using an established probabilistic anatomical cortical atlas (Desikan et al., 2006). Source inversion methods within SPM are based on Minimum Norm assumptions, whereby the MSP method generates source estimates that optimise scoring of covariance matrices (model evidences) and minimize error in localization estimates. For each epoch of data, cortical time series was calculated for each of the 68 regions and was based on source estimates to the vertex closest to the centroid with a minimum Euclidean difference criterion (VOI radius of 5 mm).

Functional connectivity was next measured across all reconstructed cortical parcellations by measuring phase synchronization using the debiased weighted phase lag index (dwPLI) approach. This metric has several advantages as it captures: (i) orthogonalized estimates of the imaginary of coherency between pairs of signals; (ii) minimized volume conduction effects generated by EEG via the exclusion of zero phase lag connections (Vinck et al., 2011) and (iii) zero lag correlations are likely to yield more reliable estimates of network connectivity (Lai et al., 2018). We thus used dwPLI to assess measure phase lag correlations across four oscillatory frequency bands: (i) theta (3.5 – 7.5 Hz), (ii) alpha (8 – 15 Hz), (iii) beta (16 – 31 Hz), and (iv) gamma (32 – 61 Hz). These oscillatory bands were chosen on balance with similar studies analyzing oscillatory activity in these frequency bands across PD cohorts. Additionally, power spectral analyses (via Pwelch’s estimate) were performed in both PD and HC groups at rest and task states (see Supplementary Figure 1). These power spectral plots indicated that electrophysiological data collected was within the frequency ranges described (Cozac et al., 2016; Hassan et al., 2017). Using our 68 regional time-series, we measured dwPLI between pairs of orthogonalized time series (i.e. phase lags between amplitude envelopes computed via the Hilbert transform). Phase lag correlations were averaged between two estimates derived in both directions (signal X relative to Y, and vice versa). All combinations of cortical time-series in each specific oscillatory band were organized by experiment condition for each subject (with connectivity graphs comprising 68 nodes) and normalized via the fisher-Z transform to parametrically align datasets. We next grouped these weighted, unthresholded, and undirected connectivity matrices into: (a) PD patients and (b) healthy control (HC) groups, for further assessment via a network-based statistics (NBS) approach (Zalesky et al., 2010). This approach is akin to neuroimaging voxel-based analyses where cluster-level thresholding of statistical parametric maps are accounted for by multiple comparisons testing and correcting for numerous univariate tests of connectivity. NBS enables us to determine differences in network topologies related to the experimental condition and group contrasts. We used 50,000 permutations to ascribe statistical significance at *p*<0.05 (family wise error, FWE network correction). Using NBS, we first identified group-level connectivity differences in each oscillatory band between PD patients and matched controls. Here, between group comparisons via unpaired t-tests were performed for each frequency band (e.g. PD theta vs HC theta) to test the hypothesis that there would be increases in connectivity (PD > HC), with alternative hypothesis testing for decreased connectivity (PD<HC), at both rest and task states. Further, NBS was used to identify pairwise functional connectivity differences within-groups at rest and during the task to examine rest-task interaction effects. For this we performed paired t-tests where only components that survived a network-level threshold of *p*_FWE_ < 0.05 were declared significant. Our paired t-tests enabled us to identify key functional connectivity changes in all oscillatory activity interacting between rest and task conditions, providing supporting results to the group-level graph analyses. Significant edges in frontal, parietal, temporal and occipital networks as identified by all NBS contrasts were visualized by connectograms in each experimental condition (http://immersive.erc.monash.edu.au/neuromarvl/).

We next interrelated symptom-specific measures with our connectivity metrics across networks via a canonical correlation analysis (CCA). CCA provides a robust, multivariate framework in which maximal covariations (also referred to as canonical variates) between two sets of variables can be determined. Investigating linear combinations between two sets of distinct variables (e.g. electrophysiological and clinical symptoms) offer insights into positive-negative interrelationships of brain-based data with clinical data. This single, integrated, multivariate method of analysis (Smith et al., 2015) reduces dimensionality of numerous brain variables and offers a more generalized view into common patterns of variation that exist between two orthogonal sets of data features. In our case, CCA seeks to explicate the interrelationship of brain connectivity networks with PD symptom measures. CCA ascribes positive and negative weights of connectivity in segregated brain networks and common symptom profiles to determine the association between these sets of variables in PD. To provide CCA the appropriate inputs, we were first guided by our NBS contrasts in identifying key fronto-temporo-parietal networks involved in rest and task-states. Based on these contrasts we next derived median within-network connectivity values, for each subject and each condition, in segregated brain networks. Median connectivity values were also derived between-networks. We next reduced the dimensionality of brain network variables to mitigate for overfitting effects for the CCA analysis. Here, including all within and between network variables for CCA is not viable, as it increases the likelihood of spurious correlations by chance which could conflate interpretations for any significant brain network and symptom relationships. In this regard, a principal component analysis (PCA) was applied to median connectivity values to examine whether rest or task states best explained within and between brain network response, so that an optimal number of brain network variables were included in our CCA to account for overfitting effects. CCA testing was controlled for family-wise error correction (*p*_FWE_ < 0.05) via permutation testing (15,000 permutations) to determine whether results reject the null hypothesis (Lin et al., 2018). Null CCAs represent random permutations of maximal correlations across subjects, corrected for multiple comparisons, wherein statistically significant correlations (*p*_FWE_ < 0.05) are compared against null distribution values. Once a significant mode of covariation between brain and symptoms data was identified, family-wise error corrected confidence intervals were then derived based on a bootstrapping procedure (5000 resamples), to ascribe CCA weights of brain network and symptoms. These canonical weights are calculated and represented in a single integrated, positive/negative axis view. For CCA analysis, additional tests were performed to ensure (1) exclusion of any subjects with missing symptom measures for dataset completeness, (2) outlier data and non-linear trends did not affect CCA outputs, and (3) multicollinearities or singularities amongst variables were acknowledged and/or minimized. Regarding the latter point, HAM-A and HAM-D scores in our PD cohort are strongly correlated with each other (r = 0.81, p < 0.001). No other significant interactions between other symptoms (including patient age) were identified. We have included these into our CCA analyses, but additionally report results of the CCA without HAM-A and/or HAM-D. Additionally, we also generated time-series surrogates using a phase randomization approach and re-performed all analysis steps following our inversion to check that phase lag correlations were not due to chance (Prichard and Theiler, 1994). To assert the validity of our results we also performed a separate train-test validation via a *k*-fold cross validation method (*k* = 10). CCA on a randomly selected “train” dataset comprising 70% of the subjects was run 10 times, and the estimated CCA network and symptom weights were then applied to “test” CCAs on subsets comprising the remaining 30% of subjects. **Figure 2** visually captures the analytic pipeline for our data.

**Figure 2:**
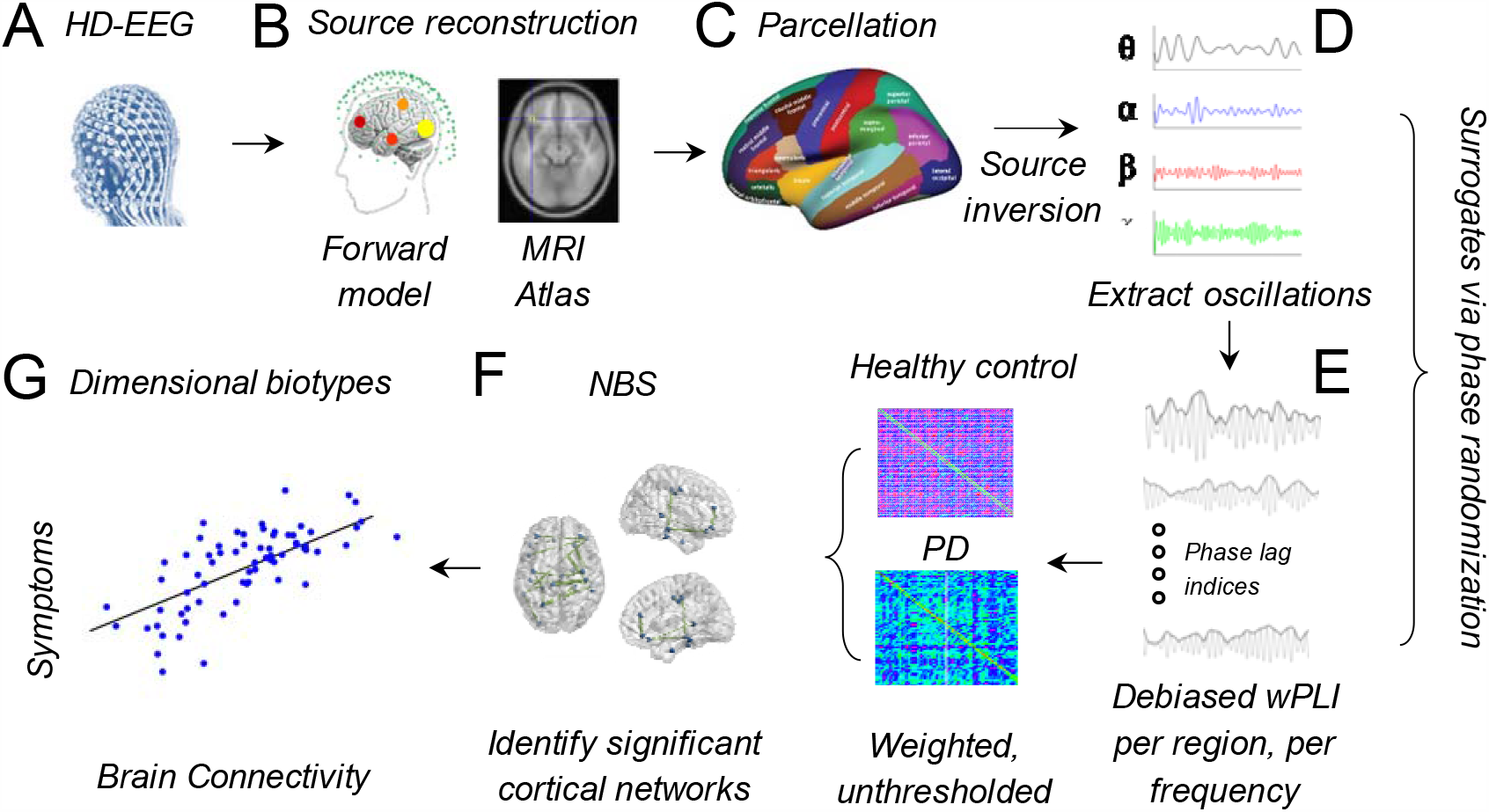
Data analysis pipeline. (A) HD-EEG during resting and task states. (B) Source reconstruction. (C) Parcellation and extraction of regional time-series. (D) Oscillatory activity bandpass filtered and (E) synchrony measures. (F) NBS to assess connectivity (G) CCA on networks and symptoms.

## 3.0 Results

### 3.1 Behavioural measures

Table 3 shows the outcomes for reaction time and accuracy for PD patients and Controls.

**Table 3:** Behavioural outcomes for Parkinson’s disease (PD) patients and Controls for each of the four conditions.

| Behavioural measure | Condition | PD (n=70) | Controls (n=21) |
| --- | --- | --- | --- |
|  |  | Mean (SD) |  |
| Reaction time (ms) | Negative-Negative | 940.34 (208.98) | 892.62 (151.07) |
|  | Negative-Neutral | 926.09 (197.83) | 888.69 (133.97) |
|  | Neutral-Negative | 965.57 (209.87) | 902.15 (142.05) |
|  | Neutral-Neutral | 912.13 (198.83) | 873.16 (115.24) |
| Accuracy<br>(%) | Negative-Negative | 75.7 (16.1) | 78.8 (16.5) |
|  | Negative-Neutral | 83.2 (11.6) | 86.3 (6.8) |
|  | Neutral-Negative | 74.4 (17.4) | 80.3 (15.9) |
|  | Neutral-Neutral | 85.7 (10.2) | 87.4 (7.0) |

#### 3.1.1. Reaction time

Reaction time data was subjected to square root transformations to normalise the distribution. Analysis was performed using the transformed data; however real time results are reported for ease of interpretation. A significant main effect of Congruency (F_1,86_=5.12, p=0.03) was observed, reflecting longer reaction times for incongruent (Mean=934.19ms, SD=179.93ms) compared to congruent (Mean=916.24ms, SD=179.41ms) prime-target pairs regardless of group. No other significant main effects or interactions with HAMD, HAM-A and LEDD were observed.

#### 3.1.2. Accuracy

Response accuracy data underwent arcsine transformations to normalise the distribution. Analysis was performed using the transformed data; however raw percentages are reported for ease of interpretation. A main effect of Target valence (F_1,86_=4.28, p=0.04) was observed, in which accuracy was lower for negative (Mean=76.1%, SD=16.3%) compared to neutral (Mean=85%, SD=9.7%) target valence trials regardless of group. No other significant main effects or interactions with HAM-D, HAM-A and LEDD were observed.

### 3.2 EEG Group level contrasts

Between group contrasts, via unpaired t-tests, indicated significantly increased theta and gamma connectivity in frontoparietal and frontotemporal networks, respectively, in PD patients compared to healthy controls, during resting-states (*p*_FWE_ < 0.05, *t*-statistic = 4.2). For task-states, PD patients had significant decreases in theta and gamma connectivity in the same networks, with specific connectivity decreases in observed in frontoparietal, frontotemporal, frontooccipital and temporo-parietal networks (*p*_FWE_ < 0.05, *t*-statistic = 3.2). No significant differences were reported in group contrasts of alpha and beta networks during rest and task states. **Figure 3** encapsulates the significant connections observed in our experimental conditions.

**Figure 3:**
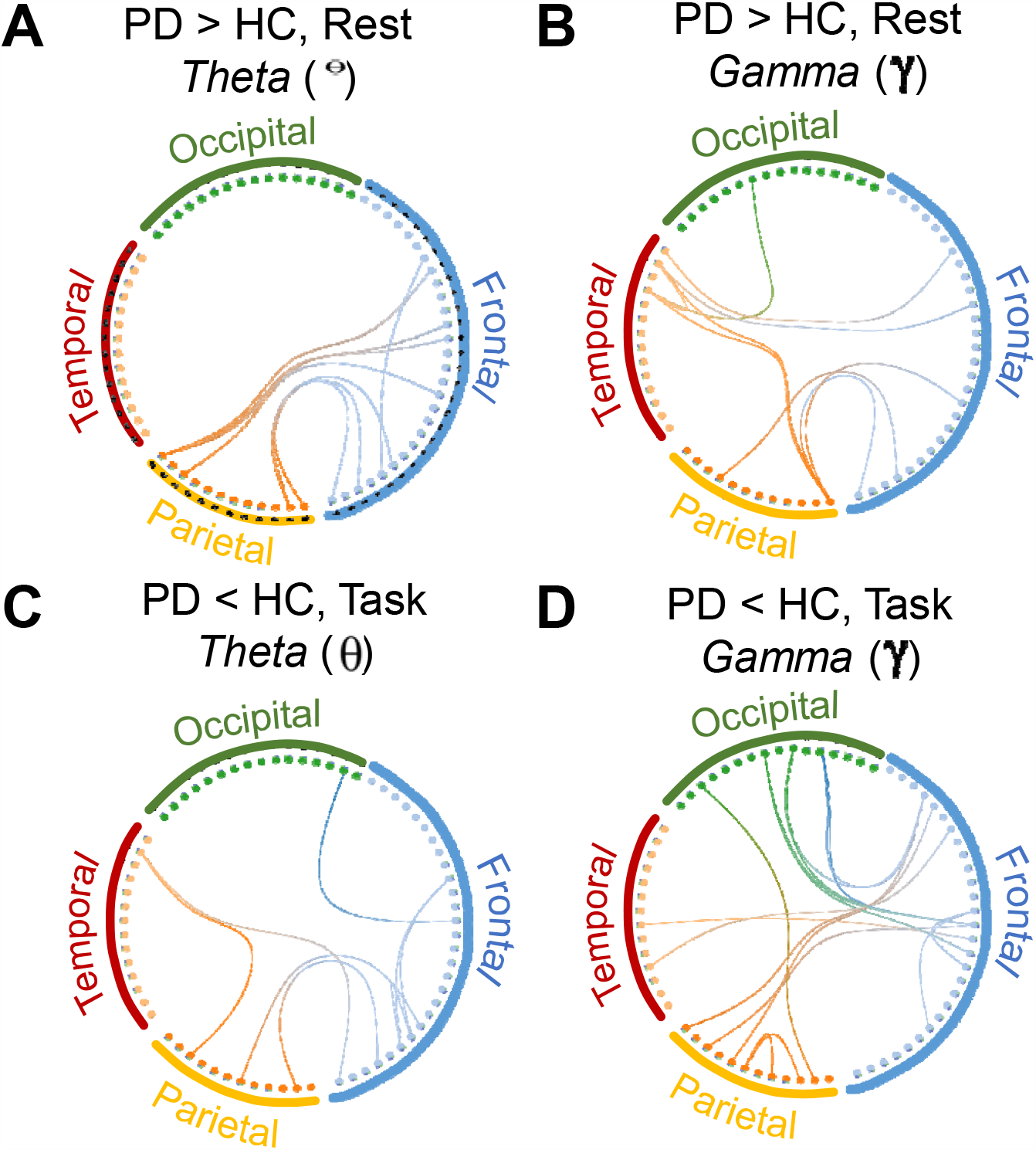
Group differences in functional connectivity as indicated by NBS. At rest, increases in connectivity within (A) theta and (B) gamma bands were observed in PD compared with HC; (C) During task-states, decreases in theta and (D) gamma connectivity were observed. For each of the plots, brain regions are demarcated by specific colors as designated by Desikan-Killany atlas coordinates: frontal = blue, parietal = yellow, temporal = red, and occipital = green. The color of each connection edge represents which “from” regions and which region it is connected “to” according to the atlas coordinates (e.g. blue connections start from Frontal regions).

### 3.3 Linked dimensions of theta and gamma connectivity with affective symptoms in PD

To optimize our CCA analyses and number of variables, dimensional reduction of between network and within network variables were performed. Here, pairwise functional connectivity contrasts further indicated key fronto-temporo-parietal network interactions between rest and task states (**Figure 4A**, *t*-statistic = 9). To further examine which between-network modes of oscillatory activity explained variance between rest and task states, a PCA on between and within theta and gamma networks in both brain states was performed. PCA outputs indicated that between network fronto-temporal and fronto-parietal connectivity values at rest was explained by gamma connectivity (85.7% and 80.2% of explained variance, in each between-network respectively) compared with theta (14.3% and 19.8% of explained variance, in each between-network respectively). PCA outputs of within networks indicated that frontal, temporal and parietal band activity was explained by theta band (Theta connectivity in frontal: 97.1%, temporal: 99.2% and parietal: 98.8%) compared with gamma (2.9%, 0.8%, 1.2%, in each within-network respectively) during task-states.

**Figure 4:**
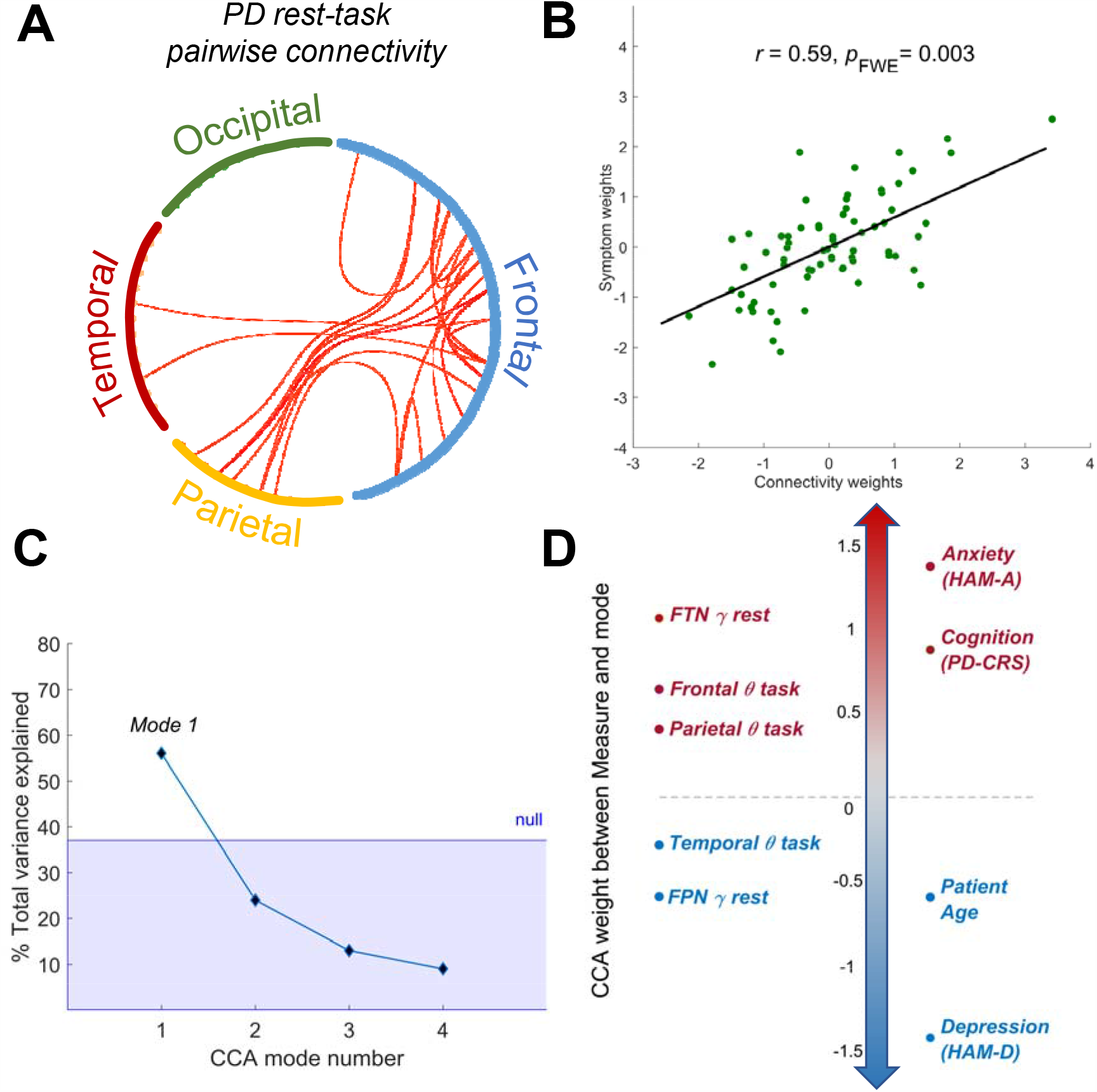
Theta and gamma connectivity is linked to affective and cognitive symptoms in PD. (A) Pairwise connectivity associated with affective priming. Here, red connection edges indicate pairwise relationships observed for theta and gamma between rest and task states. Brain regions are demarcated by specific colors according to the Desikan-Killany atlas coordinates: frontal = blue, parietal = yellow, temporal = red, and occipital = green. (B) Mode of co-variation (*p*_FWE_<0.05) in PD (C) Variance of significant CCA mode contrasted against the null CCAs, as determined via permutation testing. (D) Significant network and symptom CCA weights as determined via bootstrapping procedures.

Taken together, these networks provide focused inputs for our CCA analysis, which reveal a significant mode of co-variation linking connectivity with mood and cognitive symptoms in PD (**Figure 4B**, *r* = 0.59, *p*_FWE_ = 0.003).

CCA results were validated with surrogate time-series (see **Figure 2**) confirming the same significant mode of co-variation (*p*_FWE_ = 0.007). No significant task-specific differences to connectivity values were observed within PD groups during either incongruent or congruent conditions. Additionally, contrasts of incongruent versus congruent conditions in PD groups yielded no significant differences in connectivity values. The average correlation coefficient, via a k-fold cross validation (*k* = 10), indicated a range between *r* = 0.54 to 0.66, with a standard deviation of 0.1. Furthermore, in examining the percentage of total variance of our CCA modes, we find that the first mode explains close to 60% of the total variance in the cohort (**Figure 4C**), in contrast to our null models (median total variance of null models was 18%). The interaction between rest and task-states offers an insight on how affective and cognitive symptoms are mediated by cortical oscillatory activity in within and between circumscribed oscillatory networks (**Figure 4D**).

Here, we observed that for PD there was an increase in gamma connectivity weights between frontotemporal regions and increases in theta connectivity weights within frontal and parietal networks. Increases in these network connectivity weights corresponded with higher anxiety and cognitive impairment ratings (**Figure 4D**, highlighted in red italicized font). Conversely, a significant decrease in frontoparietal gamma network activity was followed by decreased theta connectivity within temporal networks. Decreases in these weight loadings corresponded with increased depression ratings and associated with older PD patients (**Figure 4D**, highlighted in blue italicized font). In linking these opposing changes to the overall mode of covariation, we identify a multivariate relationship in PD wherein a worsening presentation of anxiety, depression and cognitive symptoms of PD corresponded with key changes to the fronto-temporo-parietal network both during rest-states and when these brain networks are under cognitive load (task-states). Additionally, the exclusion of anxiety or depression scores from the CCA analysis did not change the overall relationship of brain network variables with symptoms (with HAM-A excluded: *r* = 0.51, *p*_FWE_ = 0.032 and with HAM-D excluded: *r =* 0.50, *p* = 0.031).

## 4.0 Discussion

Parkinson’s disease (PD) is characterised by a substantial clinical heterogeneity in affective and cognitive symptom presentation which underscores the complexity of the neurodegenerative condition (Poletti et al., 2012). Here, we investigated how anxiety, depression and cognitive symptoms in PD could be described by a multivariate relationship that links cortical network activity with their symptoms both at rest and during an affective processing state. Our study finds a novel multivariate association between functional connectivity of brain oscillations at rest and during the affective priming task, as measured by HD-EEG, with increased symptoms of anxiety, depression and impaired cognition. Specifically, connectivity increases in frontotemporal gamma, and within-network theta in frontal and parietal regions were correlated with increases in anxiety and cognitive impairment. Conversely, connectivity decreases in frontoparietal gamma, and within-temporal region theta were related to increased symptoms of depression and older PD patient age. Importantly, we find key evidence that monitoring connectivity of the fronto-temporo-parietal network changes between rest and task elucidates upon the overall severity of affective and cognitive symptoms in an ageing PD population. The study of these associations thus provides a critical advancement into the value of measuring cortical network activity in a single, multivariate construct with three highly prevalent neuropsychiatric symptoms in PD – anxiety, depression and cognitive impairment.

Whilst prior resting-state functional imaging studies in PD have identified changes in functional connectivity of the fronto-temporo-parietal networks and their association with poorer symptoms of depression and cognitive impairment, less is known about the association anxiety in PD plays in mediating brain network activity. Prior studies of resting-state fMRI in PD for example have indicated that a combination of decreases within the frontoparietal network (FPN) and default mode network (DMN), along with increases in the prefrontal cortex have been associated with increases in cognitive impairment (Amboni et al., 2015; Disbrow et al., 2014; Li et al., 2018) and higher rates of depression (Sheng et al., 2014). It has also been previously found that slowing of oscillatory activity in lower frequencies, such as theta power at rest, is a notable feature of PD progression (Dubbelink et al., 2013; Shine et al., 2014; Stoffers et al., 2007), and potentially indicative of early cognitive decline. Our group level contrasts build upon these general observations at rest in increased theta connectivity (**Figure 3A**) of the frontoparietal networks in PD patients. Moreover, the opposing connectivity changes at rest (as highlighted by the CCA, **Figure 4D**) of frontotemporal and frontoparietal gamma could suggest the potential role of activity arising from the insula and reduced prefrontal-striatal connectivity in those patients where increases in anxiety, depression and cognitive impairment have been observed (Christopher et al., 2014; Oosterwijk et al., 2018). This could also explain why decreases in frontotemporal connectivity are present in lower frequencies, as previously observed in those patients with cognitive deficits (e.g. alpha) (Hassan et al., 2017).

Cognitive and affective task states, have provided further insights into potential dysfunctions of executive functioning and decision-making processes underlying the severity of affective and cognitive symptoms in PD. Our observation of theta connectivity increases during task-states of within-network theta in frontal and parietal areas could relate more broadly to general aspects of cognitive control (e.g. conflict) and its relationship to heightened anxiety states (Cavanagh and Frank, 2014; Cavanagh and Shackman, 2015). This proposal is consistent with the notion of dopaminergic depletion of the fronto-striatal macrocircuits in PD which in turn impact affective decision-making processes (Bell et al., 2015). Associations between anxiety and cognitive impairment in PD is described in a recent clinical study of newly diagnosed PD (Dissanayaka et al., 2017b). Increased theta oscillatory connectivity in both anxiety and cognitive impairment may suggest a shared network between these two complex neuropsychiatric manifestations in PD, and warrants further investigations in future.

Our result suggesting that decreases in temporal region theta oscillations is linked to depression symptoms is in line with a previous fMRI study where decreases in temporal network connectivity and increased depression have been found in PD (Lou et al., 2015). This may reflect lower semantic activations in the temporal lobe during depressive states in PD, which is relevant considering our affective priming task utilizes words with emotional valences. The relationship between anxiety, cognitive impairment and affective-states brain networks in PD is however still emerging (Oosterwijk et al., 2018). Emotion valence studies have also found that reduced activity in the superior temporal gyrus is associated with cognitive function, which could tie into aspects of temporal lobe hypometabolism in PD patients (Bell et al., 2017; Sieger et al., 2015). Taken together, the transitions in cortical connectivity networks observed between rest and cognitive-emotion task-states highlight the key role theta and gamma plays in mediating effects of anxiety, cognitive impairment and depression severity in PD.

The interaction of resting and task-state brain networks are, in general, more well-established in examining the effects of depression and age, both in PD (Hu et al., 2015; Lou et al., 2015) and non-PD populations (Alexopoulos et al., 2012; Schultz et al., 2017). Although our study focuses on a non-demented PD sample, we posit that the increasing presentation of affective and cognitive symptoms in PD are reflected by specific cortical network interactions that are readily delineated during transitions between resting and cognitive states. This is evident in our CCA, where older PD patients and higher depression ratings have decreased connectivity in the frontoparietal network at rest and temporal regions during task-states. Therefore, we must consider the association of depression and increasing patient age, where examination of cortical activity within the fronto-temporo-parietal networks could highlight dysfunctions in decision-making during response to affective content and overall processes of executive functioning.

Findings from the present study reveal that the mode of covariation between rest and task-states of functional connectivity with clinical symptoms of depression, anxiety, and cognition in PD are defined by a multivariate association.

### 4.1 Limitations

There are some limitations to our study which could be explored in future investigations. First, given the heterogeneity of affective and cognitive symptoms in PD, factors such as an age, dosage of neuropsychiatric medication, and overall degree of motor impairment during the progression of disease need to be considered. This could be addressed via larger and broader PD cohorts which can facilitate more sensitive measures of brain-symptom relationships (including hierarchical clustering approaches), wherein subtypes of mild cognitive impairment (MCI), depression and anxiety can be further probed. Secondly, our study primarily focused on a cohort where dopaminergic medication was optimally functioning for each PD patient. Future investigations on the effect of dopaminergic “off” states need to be considered to understand the effect of fluctuating affective and cognitive symptoms and its relation to the stability of cortical oscillatory activity. As it is generally not practical to obtain drug naïve patients with PD, conducting a study with a drug naïve patient group will prohibitively increase the study duration and its associated costs. Thirdly, to avoid having a complicated paradigm we included negative vs neutral words, and did not include positive stimuli. Fourthly, the relative sample size difference between PD and HC groups, and the absence of an HC group with elevated anxiety, depression and/or cognitive impairment scores limits our ability to assert whether network changes are specifically related to affective symptoms found in PD cohorts or expected relationships for individuals with elevated affective symptoms. Though our study is limited by these factors, our comparison with HC is mainly used to track network related differences (i.e. NBS contrasts) occurring at rest and during task states, prior to CCA analyses of PD groups. In this regard, network related differences could also suggest generalized effects that occur during resting and cognitive states, that are independent of PD neuropathology. Future approaches would benefit from similar sample size and phenotypes for between-group comparisons to closely explain electrophysiological differences observed in affective states present in both PD and non-PD groups. Finally, the relationship between cortical connectivity with affective and cognitive symptoms in PD potentially highlights the intrinsic coupling that exists between theta and gamma (Canolty et al., 2006; Lisman and Jensen, 2013), which could underscore the importance of these canonical oscillatory frequencies in influencing the overall severity of depression, anxiety and cognitive impairment.

## 5.0 Conclusions

To conclude, our study highlights the multivariate links between brain activity and symptoms in PD and the potential advantage of characterizing brain network transitions between resting and cognitive states (e.g. cognition and emotion). Such insights could facilitate an optimised profiling of common mood and cognitive symptoms in PD towards targeted interventions, and consideration towards the performance of brain networks in different settings which could be selectively modulated by therapy and/or modified by medication to alleviate the presence of these symptoms in PD.

## Data Availability

Due to the sensitive nature of the data obtained, patients were assured that raw data would remain confidential and would not be shared

## Acknowledgements

We thank the Royal Brisbane & Woman’s Hospital (RBWH), RBWH Foundation (RBWF) and Parkinson’s Queensland Inc for funding the Neuropsychiatry in Parkinson’s Disease Research program via the competitive research grant schemes. We thank A/Prof John O’Sullivan and principal investigators of the QPP, Prof George Mellick, and Prof Peter Silburn for assisting with participant recruitment and assisting with grant funding. We thank Prof Gerard Byrne and Dr Rodney Marsh for their general advice and assisting with obtaining grant funding from the above competitive grant schemes. We thank Dr Luca Cocchi for technical inputs on the analyses and interpretation of findings. We also thank research assistants, Alicia Rawlings and Tara Spokes for EEG data acquisition, and all participants of the study. Dr Nadeeka Dissanayaka is funded by the Lions Medical Research Foundation for Parkinson’s disease research and NHMRC Boosting Dementia Research Leadership Fellowship APP1137339.

